# Exploring Impacts to COVID-19 Herd Immunity Thresholds Under Demographic Heterogeneity that Lowers Vaccine Effectiveness

**DOI:** 10.1101/2022.07.18.22277763

**Authors:** Chloé Flore Paris, Julie Allison Spencer, Lauren A. Castro, Sara Y. Del Valle

**Affiliations:** Los Alamos National Laboratory, Information Systems and Modeling, NM87545, USA; Intelligence Community Postdoctoral Research Fellowship Program, Los Alamos National Laboratory, NM87545, USA

## Abstract

The COVID-19 pandemic has caused severe health, economic, and societal impacts across the globe. Although highly efficacious vaccines were developed at an unprecedented rate, the heterogeneity in vaccinated populations has reduced the ability to achieve herd immunity. Specifically, as of Spring 2022, the 0–4 year-old population is still unable to be vaccinated and vaccination rates across 5-11 year olds are low. Additionally, vaccine hesitancy for older populations has further stalled efforts to reach herd immunity thresholds. This heterogeneous vaccine landscape increases the challenge of anticipating disease spread in a population. We developed an age-structured Susceptible-Infectious-Recovered-type mathematical model to investigate the impacts of unvaccinated subpopulations on herd immunity. The model considers two types of undervaccination - age-related and behavior-related - by incorporating four age groups based on available FDA-approved vaccines. The model accounts for two different types of vaccines, mRNA (e.g., Pfizer, Moderna) and vector (e.g., Johnson and Johnson), as well as their effectiveness. Our goal is to analyze different scenarios to quantify which subpopulations and vaccine characteristics (e.g., rate or efficacy) most impact infection levels in the United States, using the state of New Mexico as an example.

## 1 Introduction

The first cases of a coronavirus new to the human immune system, SARS-CoV-2, were reported in December of 2019 [12]. On March 11, 2020, the World Health Organization declared that COVID-19, the disease caused by SARS-CoV-2, was a global pandemic [35]. As of June 10, 2022, COVID-19 has caused over 539 million human cases and 6.3 million deaths worldwide [36]. In the United States alone, there have been over 87 million cases and over one million deaths [36]. The State of New Mexico has sustained 543,882 cases, representing about one fourth of the population, and 7,866 deaths [23]. The impact of COVID-19 has extended beyond infections and deaths; during the peaks, the stress on healthcare systems has compromised providers’ ability to care for patients presenting with conditions other than COVID-19 [10].

Vaccine development began as soon as the first sequence data of SARS-CoV-2 were available [15], in the face of several unprecedented challenges:

1. There was a need for speed and rapid production scaling [19].
2. Optimal randomized, controlled human trial designs had to be informed by ethical considerations [26].
3. Because of the novelty of this zoonosis and the lack of previous coronavirus vaccines, there were unknowns regarding the amount of neutralizing antibodies needed, as well as possibilities for immune system cross-reactivity, including antibody mediated-disease enhancement [27].
4. Vaccine manufacturers faced difficulties intrinsic to equitable, global distribution [13].
5. Vaccine hesitancy was gaining a foothold in a variety of groups [28].

In this challenging environment, against the backdrop of a previous average vaccine development time of over ten years (Figure 1), virologists collaborated globally to make safe, effective SARS-CoV-2 vaccinations available to the public in a record-breaking 12 months.

**Figure 1:**
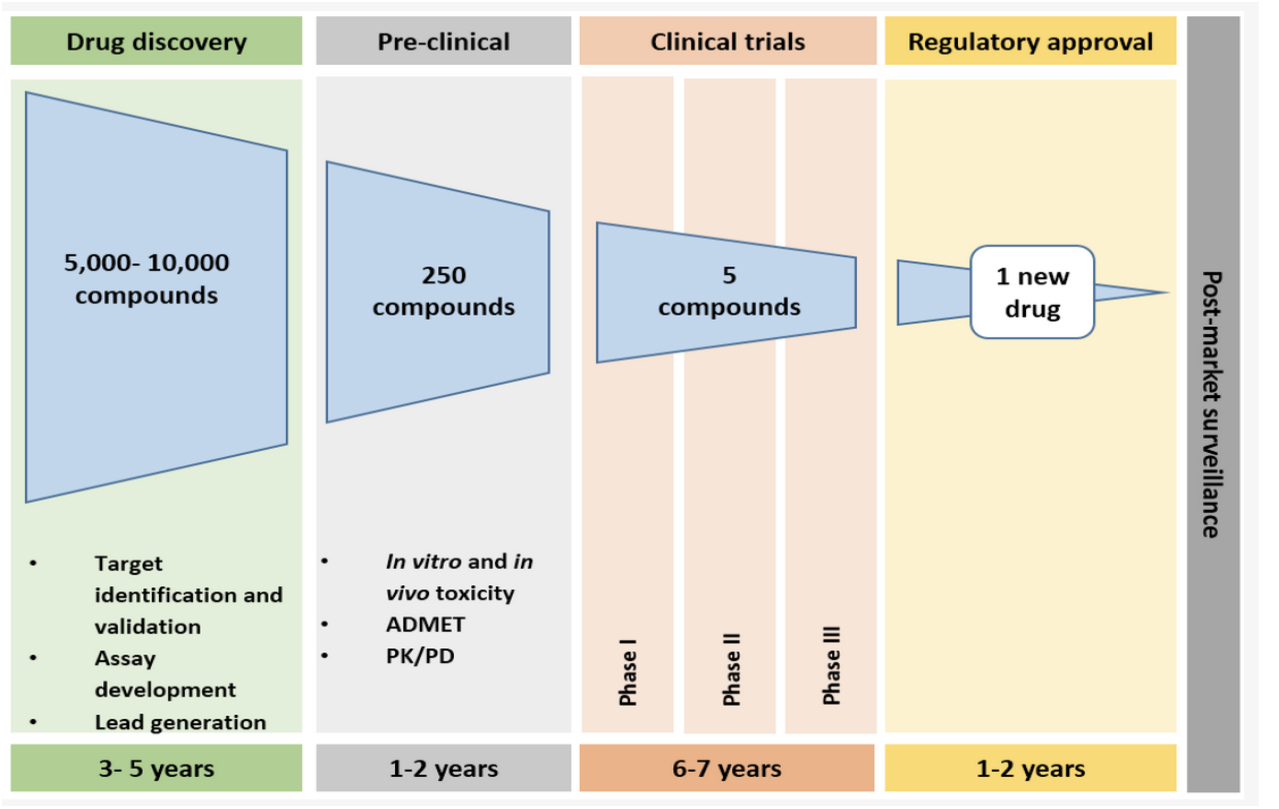
This figure illustrates the complexity of the typical vaccine development timeline [21].

**Figure 2:**
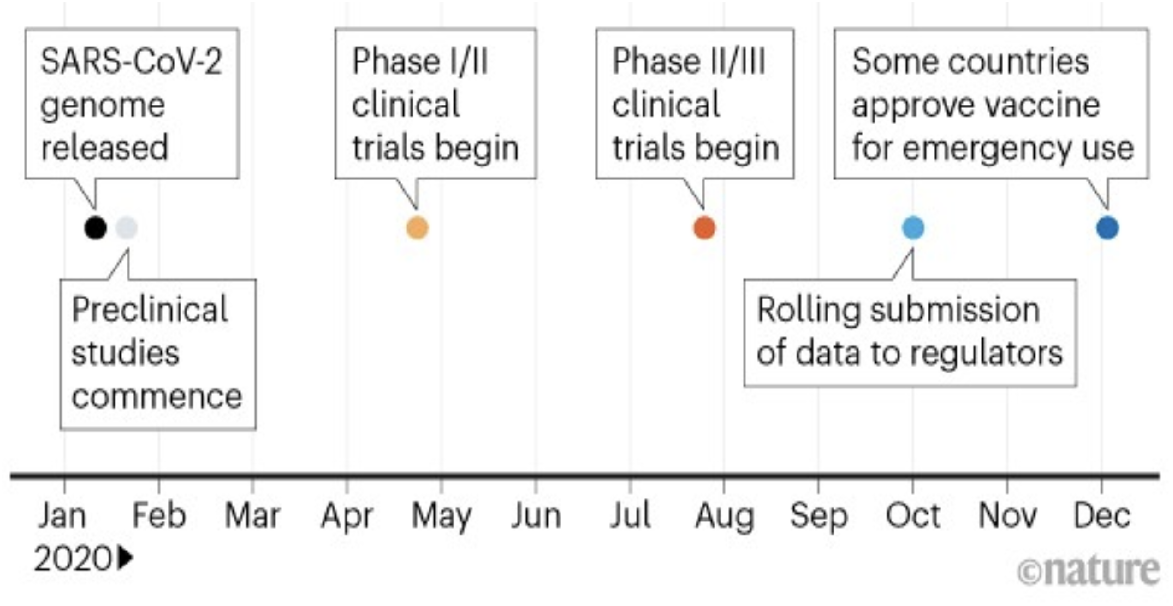
The first SARS-CoV-2 vaccine available to the public was developed jointly by Pfizer and BioNTech [7].

Of available vaccine platforms, mRNA-based and non-replicating viral vector technologies emerged as potentially the fastest and most stable, respectively [16]. The mRNA vaccine in particular could be synthetically replicated, without the need for laboratory cultures [19]. An mRNA vaccine manufactured by Pfizer-BioNTech has been available for individuals aged *≥* 16 since December 11, 2020 [25].

A second mRNA vaccine, manufactured by Moderna, has been available for individuals aged ≥ 18 since December 18, 2020 [24]. On February 27, 2021, a viral vector vaccine became available, manufactured by Janssen (Johnson & Johnson) [34].

With the availability of vaccines in the public, in the absence of eradication, an open question has been how to define a quantitative threshold for worldwide vaccine success [14]. Several studies have addressed the question of vaccine effectiveness using age-structured models, with the goal of prioritizing vaccine allocation [2, 11, 29, 37, 38]. However, age-structured modeling studies that evaluate the impacts of differing vaccine efficacies on infection levels have not been as numerous [20]. Our goal in this study was to analyze different scenarios to quantify which subpopulations and vaccine characteristics most impact infection levels in the United States, using the state of New Mexico as an example.

## 2 Methodology

### 2.1 Model Description

To model COVID-19 transmission, we use the well-known Susceptible-Infectious-Recovered mathematical model of disease transmission and expand it by adding two vaccinated populations within age-structured subpopulations. We build in four subpopulations who can cross-infect each other to construct an age-structured model. The four age groups include: ages 0-4 (group 1), ages 5-11 (group 2), ages 12-15 (group 3) and ages 16+ (group 4). The groups were chosen according to the FDA-approved vaccines per age group ([33] [32] [31]).

There are a total of six compartments for each age group: susceptible (*S*^*i*^), vaccinated by Pfizer or Moderna vaccine 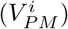, vaccinated by Johnson and Johnson vaccine 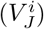, exposed (*E*^*i*^), infected (*I*^*i*^), and recovered (*R*^*i*^) (Table 1). An individual is considered vaccinated once they have completed the vaccination series (2 doses for Pfizer and Moderna, 1 dose for Johnson and Johnson). We account for the different vaccine effectivenesses by separating the two different types of vaccines, mRNA (e.g., Pfizer, Moderna) and vector (e.g., Johnson and Johnson) and letting a proportion of vaccinated individuals get exposed to the COVID-19 virus (breakthrough infections). This results in a system of 24 ordinary differential equations (Eq. 1 and Figure 3). All parameter values used can be found in Appendix A.

**Table 1:** Descriptions of State Variables

| Variable | Description |
| --- | --- |
| $S^i$ | Number of Susceptible individuals |
| $V_{PM}^i$ | Number of Vaccinated individuals (Pfizer/Moderna) |
| $V_J^i$ | Number of Vaccinated individuals (Johnson and Johnson) |
| $E^i$ | Number of Exposed individuals |
| $I^i$ | Number of Infectious individuals |
| $R^i$ | Number of Recovered individuals |

**Figure 3:**
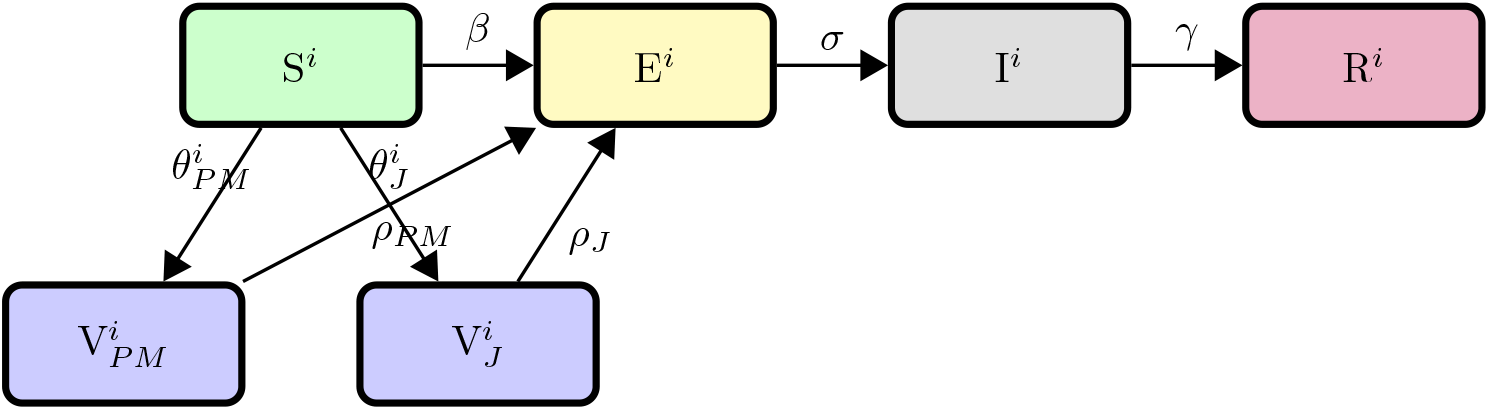
Transfer diagram for COVID-19 age-structured model.

We assume that each age group has a unique transmission factor. Thus, the transmission within each group is the same across all individuals in the group, however, it is different across the other age groups. We assume that recovered individuals do not incur waning immunity, and thus will not be reinfected within the modeled period of eight months. Vaccinated individuals within the vaccine’s efficacy umbrella are fully protected from infection for eight months; vaccinated individuals with breakthrough infections will not be reinfected after recovery. We assume that neither vaccinated individuals within the efficacy umbrella nor recovered individuals transmit the virus, however, vaccinated individuals can become infected and move to the exposed class.

Our model is homogeneous (aside from the inherent heterogeneity that the age-structured aspect of the model creates); the risk of catching COVID-19 is higher for groups 3 and 4. We assume that there are no births/deaths, that it is a closed system geographically (no inter-state travel) and that there is no movement between age groups, that is, people do not age during the simulation period. Only susceptible individuals are vaccinated; infectious and recovered individuals are not. We assume that when an individual is vaccinated, the vaccine becomes effective immediately.

### 2.2 Model Diagram

### 2.3 State Variables

### 2.4 Model Equations and Parameter Descriptions

The equations defining our model (with n = 4 subpopulations who can cross-infect each other) are given by

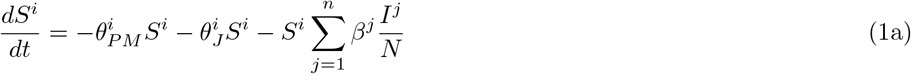

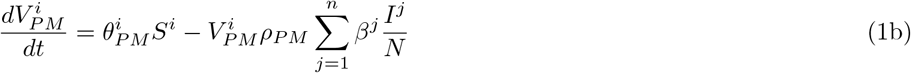

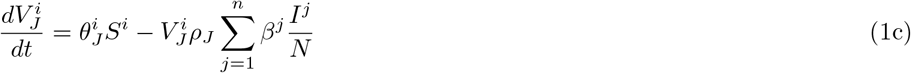

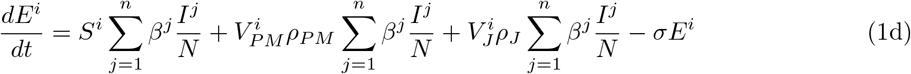

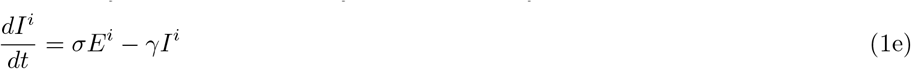

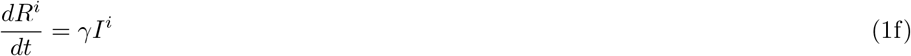

The total population is 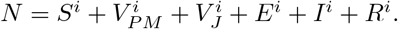

### 2.5 Estimating The Transmission Term

It has been observed that children are less susceptible to being infected with the SARS-CoV-2 virus than adults [8]. The same study found that the susceptibility of children to infection by the SARS-CoV-2 virus is 43% of the susceptibility of adults, where children are individuals 7-19 years old and adults are 20+ [8]. We use these percentages to guide our estimations of our transmission terms (*β*^1^, *β*^2^, *β*^3^, *β*^4^ in Table [2]), making the assumption that the transmission term for age groups 3 and 4 (12+) will be higher than the transmission term for age group 1 and age group 2 (11 and under).

**Table 2:** Descriptions and dimensions for parameters.

| Parameter | Description | Dimension |
| --- | --- | --- |
| $\theta_{PM}^i$ | Vaccination rate for Pfizer/Moderna | $day^{-1}$ |
| $\theta_J^i$ | Vaccination rate for Johnson and Johnson | $day^{-1}$ |
| $\beta^j$ | Rate of infection | $day^{-1}$ |
| $\sigma$ | Incubation rate | $day^{-1}$ |
| $\gamma$ | Recovery rate | $day^{-1}$ |
| $\rho_{PM}$ | Proportion of vaccinated individuals that become exposed (Pfizer/Moderna) | dimensionless |
| $\rho_J$ | Proportion of vaccinated individuals that become exposed (Johnson and Johnson) | dimensionless |

### 2.6 Estimating Vaccination Rates

For the purpose of analyzing our model, we used vaccination data from the New Mexico Department of Health (NMDOH) [22] and the Centers for Disease Prevention and Control (CDC) [3, 5]. To establish the initial conditions of our susceptible and vaccination populations for each age group, we used the NMDOH’s data on the percentage vaccinated within each age group by either the Pfizer/Moderna or J&J vaccines. For each age group, we assumed the initial susceptible population was equal to all remaining individuals.

We set vaccination rates using the CDC report on vaccination trends in children. First, we assume the vaccination rates for age group 4 (16+) is almost zero (with J&J lower than Pfizer/Moderna), as few individuals in that population are currently getting vaccinated since these population were the first to be vaccinated. For the week of February 23, 2022, the CDC reported that 80,000 children ages 12-17 received their initial dose while 8.4 million remained unvaccinated. Thus we use a vaccination rate of 0.001 for age group 3. Similarly, the CDC reported that 1% of children aged 5-11 got vaccinated the week of February 23, 2022 [5]. We take 1% of our susceptible population for age group 5-11 and estimate a vaccination rate of 0.001 for age group 2. As age group 1 (0-4) currently does not have a vaccine available, we set the vaccination rate as zero, except during our exploration of vaccination rates for children ages 0-4.

## 3 Results

## 3.1 The current vaccination landscape in individuals 5+ is not sufficient to confer herd immunity

In the absence of an approved vaccine in the youngest age group as of June 2022, we explore three scenarios to determine the impact of increasing vaccination rates in age groups 1 and 2. We first consider a scenario with the initial vaccinated population comprised of the three older age groups (individuals 5+), but assuming only individuals 12-15 are continuing to be vaccinated. In this setting, population herd immunity is not achieved and a significant outbreak results from the introduction of a new infection(Figure 4A, blue line). In our simulation, the outbreak lasts about 3 months, with a peak around day 75. The 16+ age group is the largest subpopulation in terms of individuals and also contributes the largest number of infections. Conversely, the 12-15 age group is the smallest subpopulation and contributes only 41,005 infections to the total. The two youngest age groups are disproportionately affected; they collectively make up ∼15% of the population, but contribute to ∼38% of the infections. With limited vaccinated individuals, 98.57% and 68.03% of the 0-4 and 5-11 subpopulations are infected respectively.

**Figure 4:**
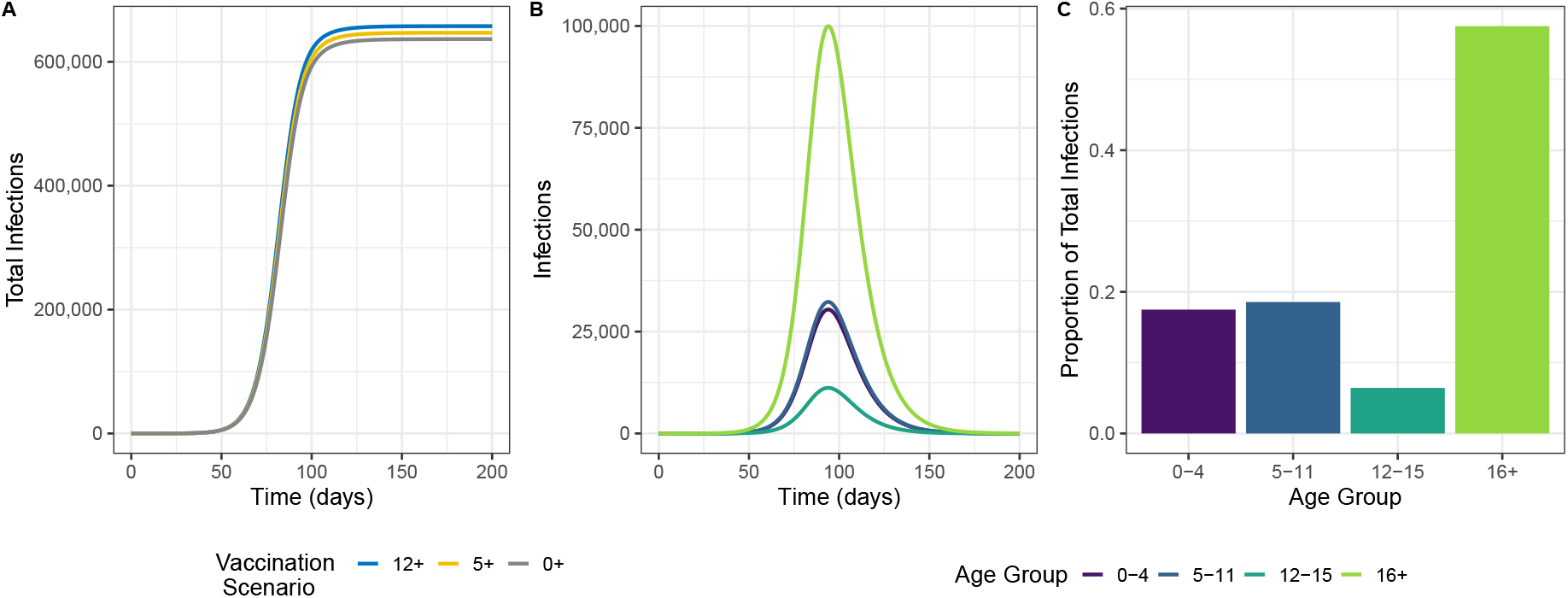
Outbreak dynamics under approximate conditions in New Mexico, spring 2022. (A)Total (cumulative) infections under three different vaccination scenarios. 12+ describes the scenario where the 5-11, 12-15, and 16+ age groups’ vaccinated populations are initialized as described in Table 3, but only the 12-15+ susceptible population continues to receive vaccinations at an appreciable rate; The 5+ scenario adds vaccination to the 5-11 age group; the 0+ adds vaccination to the 0-4 age group. (B) The prevalence of infection across the age groups for the 0+ scenario. (C) The population proportion of total infections by age group for the 0+ scenario.

Next, we find that when susceptible children in the 5-11 age group are vaccinated based on rates estimated from February 2022, herd immunity still is not achieved. At the end of the epidemic, about 30% of the total population will have been infected. The additional influx of vaccine does appreciably lower the infectious peak of age group 2, from 35,641 to 32,564 as well as the total infections. However, the impacts to other age groups are limited.

Finally, we consider a future scenario where the youngest subpopulation is vaccinated at a rate similar to that of the 5-11 age group. The vaccine confers some protection to the 0-4 age group, but the large number of unvaccinated individuals across all age groups continues to fuel an outbreak in the general population. Compared to a scenario based on realistic vaccination rates for ages 5+, approximately 10,000 fewer total individuals are infected by the end of the outbreak, representing less than a percentage of the total population.

### 3.2 Herd immunity is not possible without increases in the percentage of adults vaccinated

Given the plateauing vaccination rates in eligible age groups as of spring 2022, and potential parental hesitancy of vaccinating the youngest subpopulation when a vaccine becomes available, we explore population level gains for hypothetical increases in vaccination rates in children versus adults.

For simplicity, we combine our two younger groups into a 0-11 grouping and the two older age groups into a 12+ grouping. For each grouping, we then find increases in the Pfizer/Moderna vaccination rate, *θ*_*P M*_, that would raise the grouping’s vaccinated percentage after 250 days by increments of 5%, holding the other grouping’s vaccinated population constant at current, i.e., “baseline”, levels (see Table 3). For the younger age grouping, we consider increases through 50% since the vaccinated percentage of this combined age groups starts at 18%. We consider only increases through an additional 20% vaccinated in the 12+ age grouping since the grouping has a combined starting percent vaccinated of 78%. After finding the appropriate values of *θ*_*P M*_ while holding each group constant (row 1 and column 1 in panels A, B, C of Figure 5), we consider each unique combination. Values of *θ*_*P M*_ can be found in Table 6.

**Table 3:**
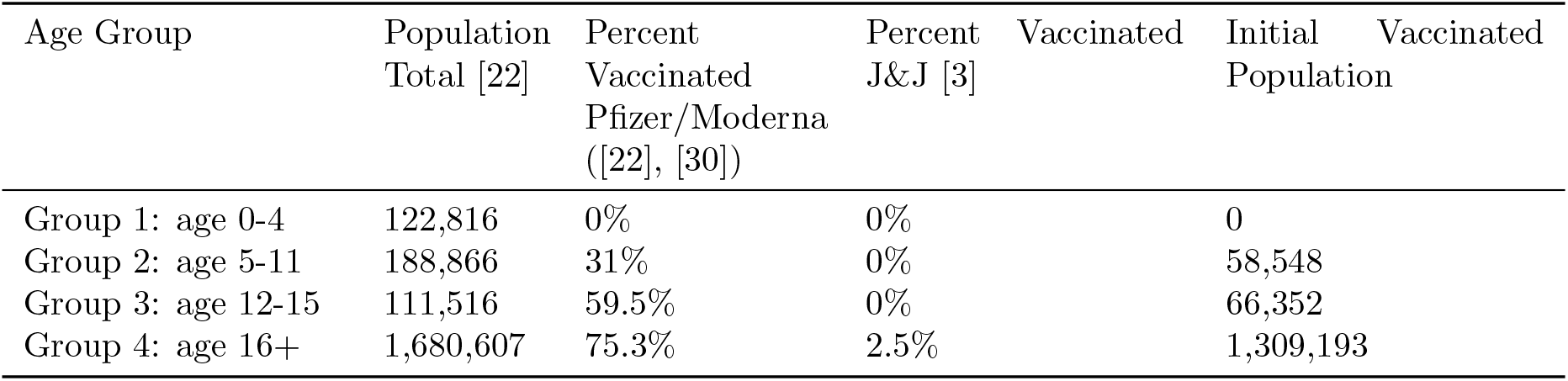
Initial conditions of population size and vaccinated subpopulations.

**Table 4:** **Comparison of percentage of total subpopulation infected with different vaccination rates**. The 12+ vaccinated simulation has rates of 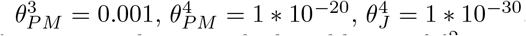. The 5+ vaccination simulation has the same rates as the 12+ simulation with the addition of 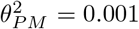. The 0+ simulation has the same rates as the 5+ simulation with the addition of 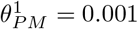.

| Age Group | 12+ Vaccinated |  | 5+ Vaccinated |  | 0+ Vaccinated |  |
| --- | --- | --- | --- | --- | --- | --- |
|  | % infected | raw infected | % infected | raw infected | % infected | raw infected |
| Age Group 1 | 98.57% | 121067.59 | 98.48% | 120958.14 | 90.60% | 111282.38 |
| Age Group 2 | 68.03% | 128487.63 | 62.61% | 118263.07 | 62.53% | 118104.03 |
| Age Group 3 | 36.82% | 41061.46 | 36.77% | 41005.889 | 36.72% | 40950.50 |
| Age Group 4 | 21.81% | 366692.16 | 21.79% | 366353.07 | 21.77% | 366009.79 |
| All Age Groups | 31.24% | 657308.85 | 30.73% | 646580.18 | 30.24% | 636346.71 |

**Figure 5:**
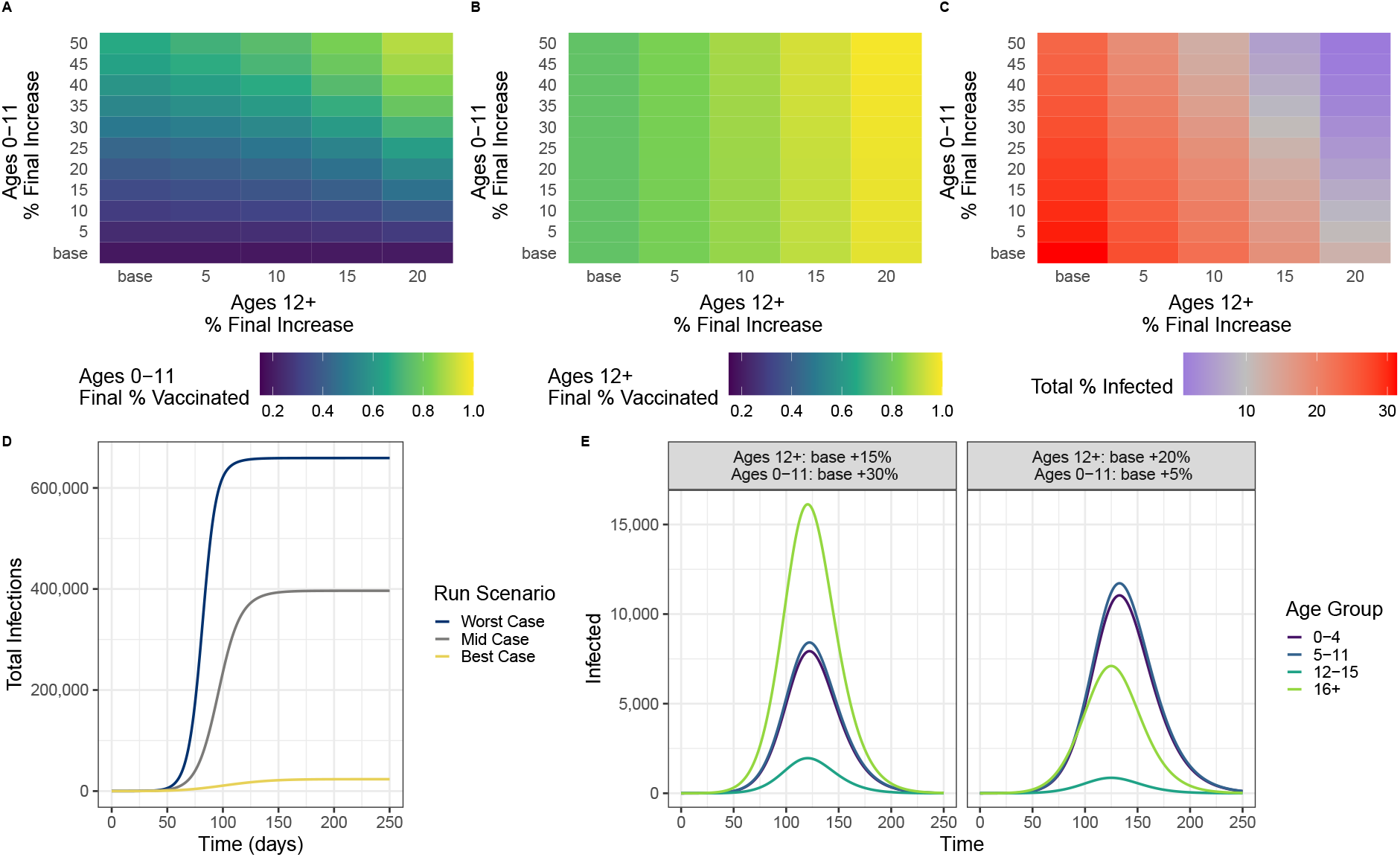
Impacts of incremental vaccination in adults versus children. In A-C, “base” refers to vaccination rates approximated from spring 2022 data for the combined age groups. (A) The final*∼* % vaccinated in the 0-11 age group after 250 days for each combination of 0-11 and 12+ increases in vaccination rates. (B) The final % vaccinated in the 12+ age group after 250 days for each combination of 0-11 and 12+ increases in vaccination rates. (C) The percentage of the population infected after 250 days for each combination of 0-11 and 12+ increases in vaccination rates. The midpoint of the scale is at 10% – a threshold chosen based on assessments of the severity of historic influenza seasons [1] (D) A comparison of cumulative infections from three scenarios. “Worst Case” represents the base case from A-C (lower left corner),“Mid Case” represents the outcome when the total % infected is 10%. In this scenario, *θ*^12+^ = 0.428 (base + 10%) and *θ*^0*−*11^ = 0.2452 (base + 50%). The “Best Case” represents the the upper right hand corner of heatmaps A-C. (E) Two scenarios that produce 10.5% of the population infected.

We find that incremental increases in vaccination rates in the 12+ grouping not only impact the percentage of vaccinated individuals in the grouping (Figure 5B), but also impact the final percentage of vaccinated individuals in the 0-11 grouping (Figure 5A). For a given vaccination rate in the 0-11 grouping (rows), the final percentage of individuals vaccinated in the 0-11 grouping rises with increases in the vaccination rate in the 12+ grouping (columns). We suspect this to be the result of fewer infections in the 12+ subpopulations that make up 85% of the total population, resulting in more individuals being able to be vaccinated before becoming infected. However, we did not see the reverse of this supplemental vaccinated effect – specifically, incremental increases in the 0-11 vaccination rate did not change the overall vaccinated percentage in the 12+ age group (Figure 5B). In Fig 5C, we see that different combinations of vaccination rates in each of the aggregated age groupings can lead to similar outcomes measured in terms of the percentage of the population infected by the end of 250 days.

Importantly, the number of infections cannot be reduced below 10% by only increasing the vaccinated proportion of the younger age groups. Reducing infection to 10% of the population is achieved either (1) with at least a 20% increase in the vaccination proportion of the 12+ grouping, to be 98% by the end of the simulation, but no changes to the 0-11 age grouping, or (2) by increasing the 12+ by 15% and the 0 − 11 population by 30% to 61%. While the end result will be similar, the size and timing of the outbreak will be impacted by the specific combination of vaccination rates. The outbreak will be slower, i.e., “flatter”, for increases in the vaccination proportion of the larger 12+ population.

### 3.3 Vaccine efficacy matters more than the rate of vaccine acquisition

For our previous analyses, we assume the vaccine to have a near 100% efficacy in preventing breakthrough infections. This assumption may have been appropriate early in the pandemic before immune-escaping variants; however, evidence from the Delta and Omicron waves in vaccinated individuals as well as from clinical studies in the 0-4 age group suggest breakthrough infections were more common as of winter 2021 [18]. Thus, we perform a sensitivity analysis to determine whether the vaccine efficacy or the rate of vaccination is more important in terms of preventing infections. We quantify vaccine efficacy by measuring the proportion of breakthrough infections, i.e., the ratio of the number of vaccinated individuals who become infected to the total number of vaccinated individuals, at the end of the simulation. We first consider changes only to the parameter *ρ* that would result in vaccination efficacies of 20-80%, where vaccination efficacy is defined as the proportion of vaccinated individuals who had a breakthrough infection (Fig 3). Values of *ρ* can be found in Table 7. Vaccination rates, *θ*, are assumed to be as they were in April 2022.

**Table 5:** Parameter values of model for three explorations

| Parameter | Value |  |  | Dimension |
| --- | --- | --- | --- | --- |
|  | Exp 3.1 | Exp 3.2 | Exp 3.3 |  |
| $\beta^{1,2}$ | 1.155 | 1.155 | 1.155 | $day^{-1}$ |
| $\beta^{3,4}$ | 1.5 | 1.5 | 1.5 | $day^{-1}$ |
| $\theta_{PM}^1$ | 0 | 0 | 0.001 | $day^{-1}$ |
| $\theta_{PM}^2$ | 0 | 0.001 | 0.001 | $day^{-1}$ |
| $\theta_{PM}^3$ | 0.001 | 0.001 | 0.001 | $day^{-1}$ |
| $\theta_{PM}^4$ | $1 * 10^{-20}$ | $1 * 10^{-20}$ | $1 * 10^{-20}$ | $day^{-1}$ |
| $\theta_J^1$ | 0 | 0 | 0 | $day^{-1}$ |
| $\theta_J^2$ | 0 | 0 | 0 | $day^{-1}$ |
| $\theta_J^3$ | 0 | 0 | 0 | $day^{-1}$ |
| $\theta_J^4$ | $1 * 10^{-30}$ | $1 * 10^{-30}$ | $1 * 10^{-30}$ | $day^{-1}$ |
| $\sigma$ [17] | 1/5.2 | 1/5.2 | 1/5.2 | $day^{-1}$ |
| $\gamma$ [6] | 1/10 | 1/10 | 1/10 | $day^{-1}$ |
| $\rho_{PM}$ [4] | $1.016 * 10^{-4}$ | $1.016 * 10^{-4}$ | $1.016 * 10^{-4}$ | dimensionless |
| $\rho_J$ [4] | $1.016 * 10^{-4}$ | $1.016 * 10^{-4}$ | $1.016 * 10^{-4}$ | dimensionless |

**Table 6:**
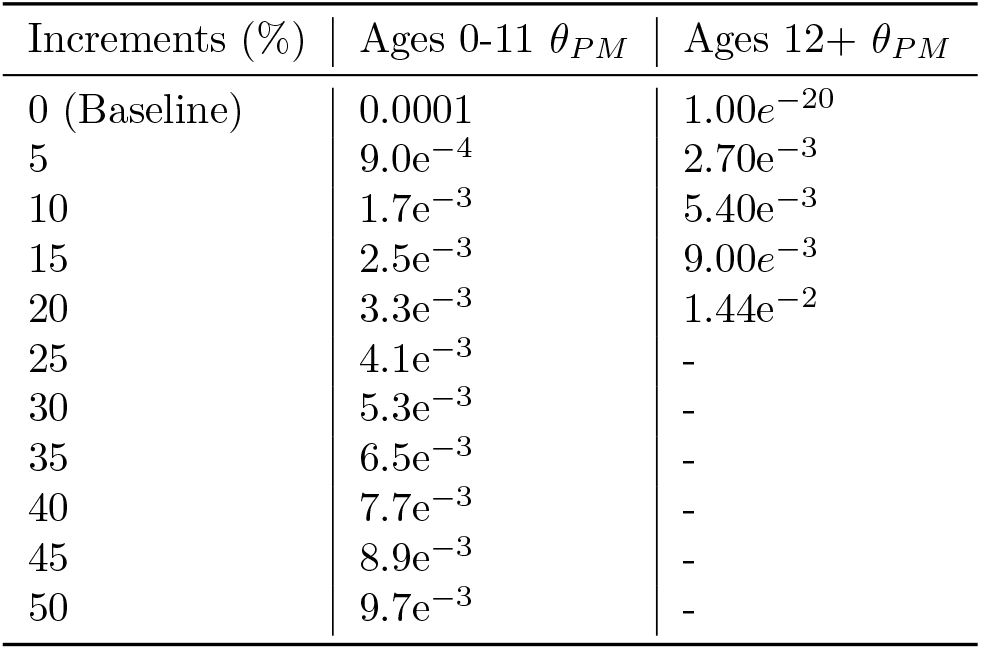
Exploratory vaccination rates for Figure 5.

**Table 7:**
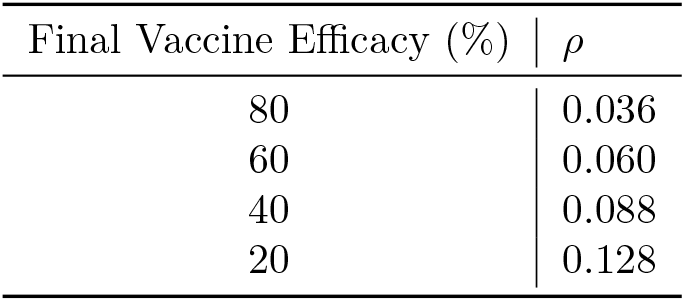
Exploratory vaccination rates for Figure 5.

Stronger vaccine efficacies results in flatter curves for all sub populations (Figure 6A). When the vaccine efficacy is low and the starting vaccinated population is high, as in age group 4, the outbreak is perpetuated by breakthrough infections, after unvaccinated individuals are depleted (Figure 6, Panel B). Because there are fewer starting vaccinated individuals in the younger age groups, the pool of completely susceptible individuals is large enough that unvaccinated individuals still make up the majority of infections. Finally, we find that total infections are more sensitive to the efficacy of the vaccine than to the rate at which they are distributed.

**Figure 6:**
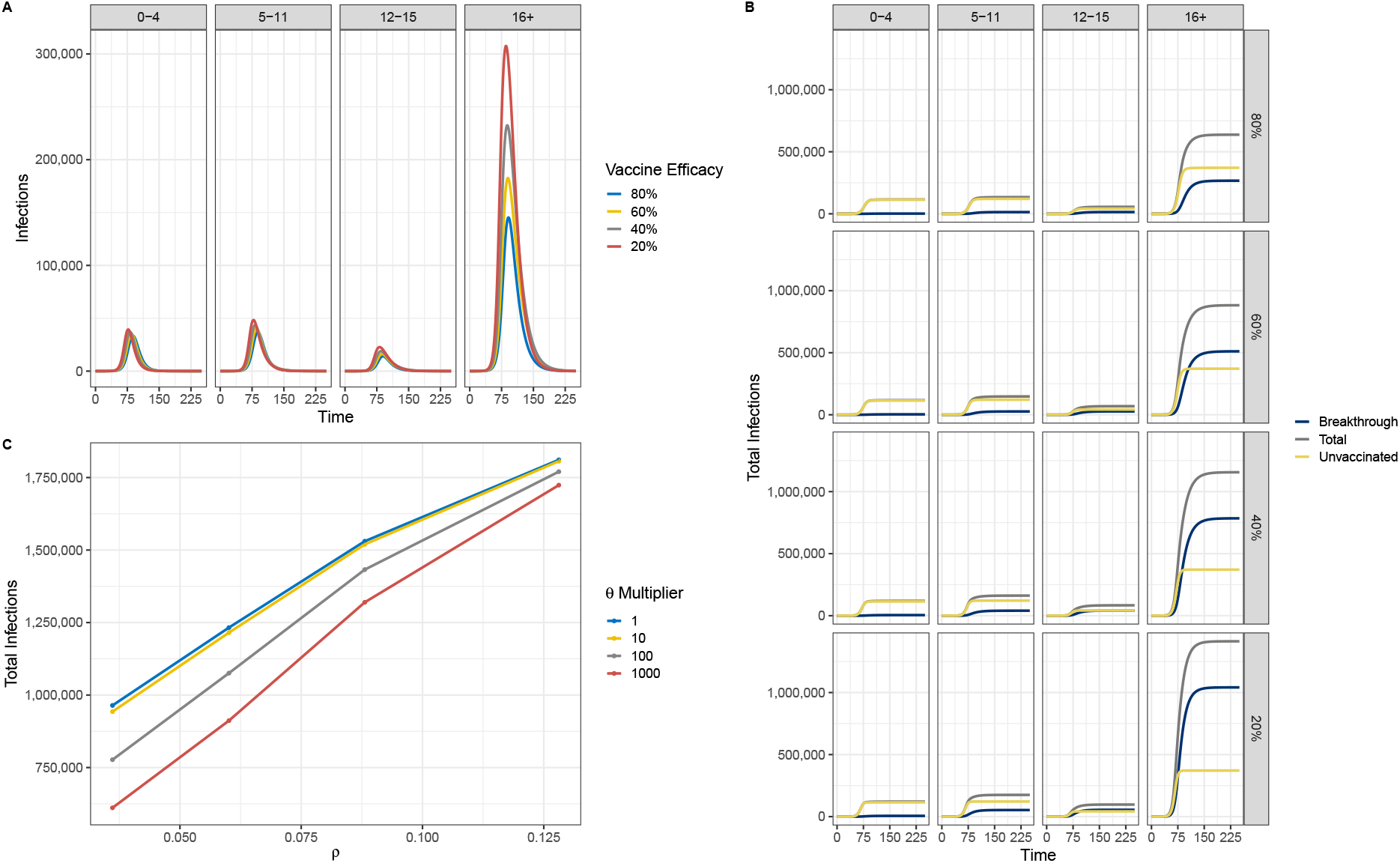
Analysis of vaccination efficacy. (A) Impacts of increasing vaccination efficacy on each of the age groups. (B) The breakdown of infections by vaccination status, age group, and vaccine efficacy. Sensitivity analysis of vaccination efficacy (as measured by *ρ* and vaccination rate, *θ*). As *ρ* increases, vaccination efficacy goes down. The *θ* multiplier denotes the multiplication factor applied to the base rates in Appendix 1 Table 5.

## 4 Discussion

COVID-19 vaccines were developed at an unprecedented rate that have significantly helped control the pandemic. Although highly efficacious, the continuing evolution of SARS-CoV-2, lack of vaccines for children under the age of 4, and vaccine hesitancy have limited their ability to halt its spread. We used an age-structured epidemiological model to examine the impact of this heterogeneity in vaccinated populations and reduced vaccine efficacy on COVID-19 outcomes.

The numerical simulation results show that in a heterogeneously vaccinated population (corresponding to 0%, 31%, 59.5%, and 75.3%), herd immunity cannot be achieved. Under this current landscape, our results show that while 16+ have the largest proportion of infections, the two youngest age groups (ages 0-4 and 5-11) are disproportionately affected, since they make up ~ 15% of the population but contribute to ~ 38% of the infections. This result supports a cocoon vaccination strategy, which consists in vaccinating persons from the immediate environment of those who might develop an illness. That is, in the absence of vaccines for children under the age of 4, family members and contacts should get vaccinated in order to protect them.

Additionally, our results showed that even if 30% of the youngest age groups were to be vaccinated, the unvaccinated population continues to fuel the spread. Thus, herd immunity cannot be achieved unless 80% of the adult population, which makes up the largest percentage of the population, is vaccinated. Finally, we showed that vaccine efficacy is more important than vaccine uptake. A highly efficacious vaccine can flatten the curve even if vaccine uptake is low.

Note that this study has several limitations:

1. Unrealistic contact patterns: Although the population was stratified into four age groups, we assumed homogeneous mixing within each age group. This assumption is unrealistic given the highly heterogeneous mixing patterns we have in the real world. However, this study is intended to provide insights and not necessarily to capture the true dynamics observed in real populations.
2. Vaccine waning: Recent studies have shown waning immunity after SARS-CoV-2 vaccination in all age groups after 6 months. However, since our simulations consider a 8-month time frame, we did not incorporate waning.
3. Vaccinated individuals cannot transmit to others: Although this assumption is typically true for other vaccines, recent evidence suggests that even vaccinated individuals are able to transmit to others. This limitation is likely to overestimate the impact of vaccine on controlling disease spread in our model in the first part of our analysis.
4. Timing between vaccine doses and boosters: In our model, we assume that people are fully vaccinated when they move into the vaccinated compartment and do not incorporate the timing required between the two vaccines as well as the 2-week period it takes for the vaccine to become fully effective. This limitation is also likely to overestimate the impact of vaccines on COVID-19 outcomes.

## Data Availability

All data produced in the present study are available upon reasonable request to the authors.

## Acknowledgements

This work is approved for distribution under LA-UR-22-2591. This research was partially funded by NIH/NIGMS under grant R01GM130668-0, which supported SYD, LC, and CP.

JAS was supported by an appointment to the Intelligence Community Postdoctoral Research Fellowship Program at Los Alamos National Laboratory administered by Oak Ridge Institute for Science and Education (ORISE) through an interagency agreement between the U.S. Department of Energy and the Office of the Director of National Intelligence (ODNI).

The findings and conclusions in this report are those of the authors and do not necessarily represent the official position of Los Alamos National Laboratory. Los Alamos National Laboratory, an affirmative action/equal opportunity employer, is managed by Triad National Security, LLC, for the National Nuclear Security Administration of the U.S. Department of Energy under contract 89233218CNA000001. The funders had no role in study design, data collection and analysis, decision to publish, or preparation of the manuscript.

## A Appendix

### A.1 Model Parameter Values

#### A.2 Basic Reproduction Number

The total infected population has eight compartments, *E*^1^, *E*^2^, *E*^3^, *E*^4^, *I*^1^, *I*^2^, *I*^3^, *I*^4^. It is important to note that R_effective_, unlike *R*_0_, does not assume a completely susceptible population but rather one that has non-pharmaceutical interventions in addition to vaccinations. We derive *R*_*effective*_ using the next generation matrix method [9]. The F and V matrices are shown below,

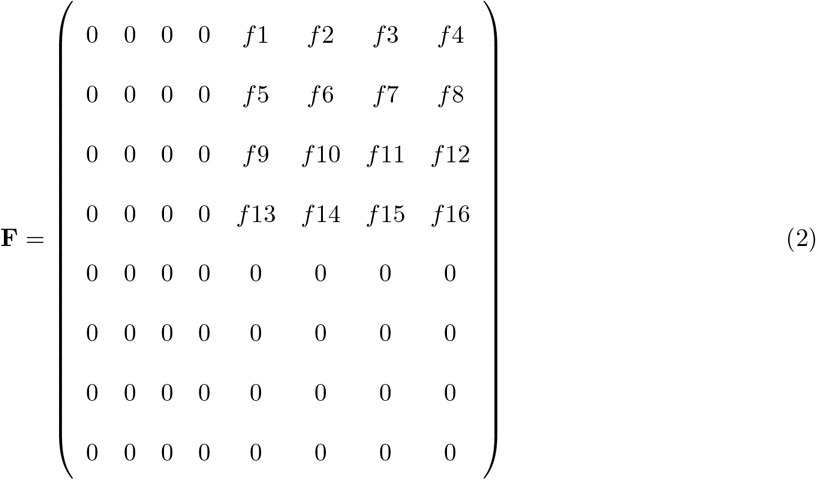

where the entries in the matrix correspond to

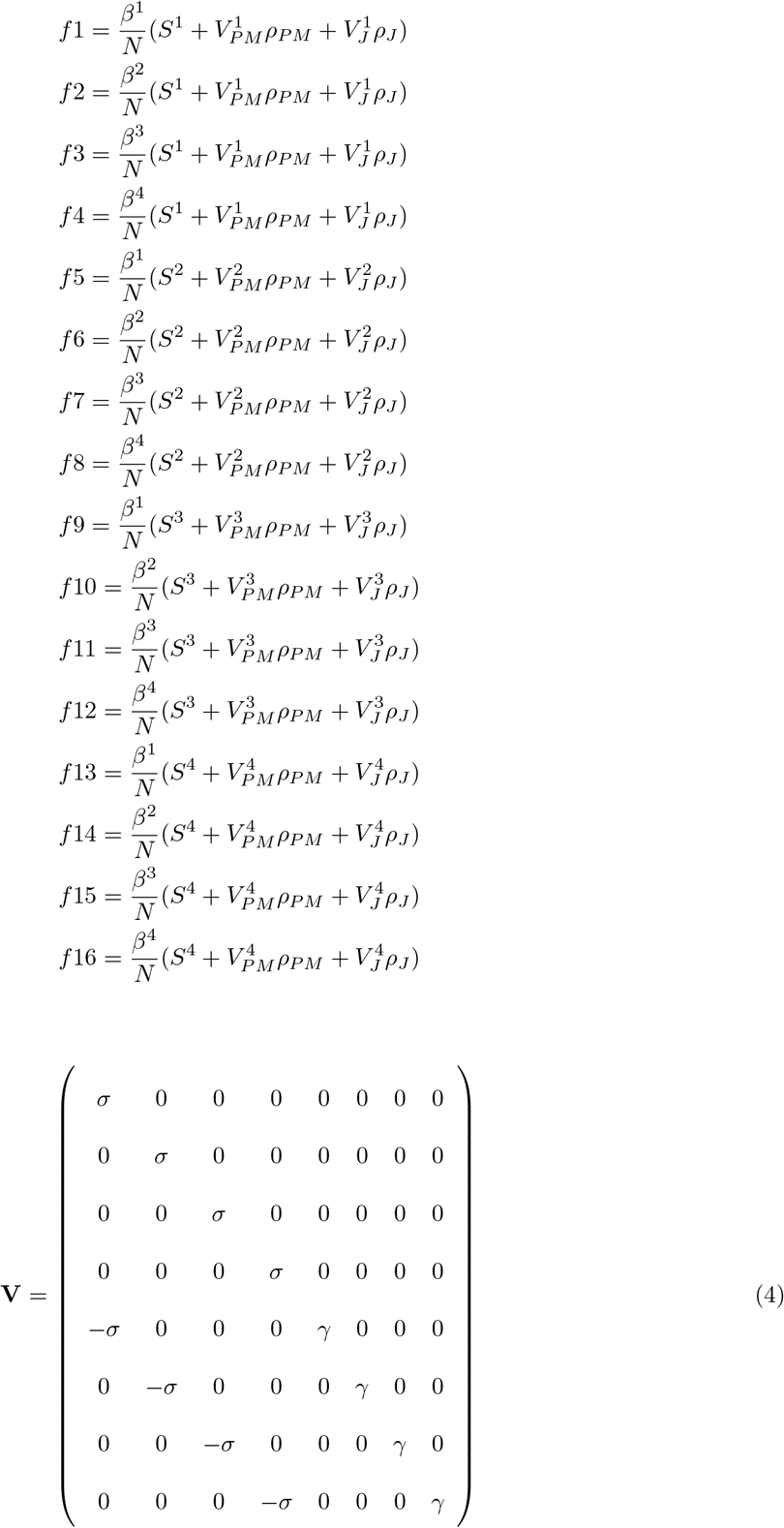

The eigenvalues of the next generation matrix (*NGM* = *FV* ^*−*1^) are

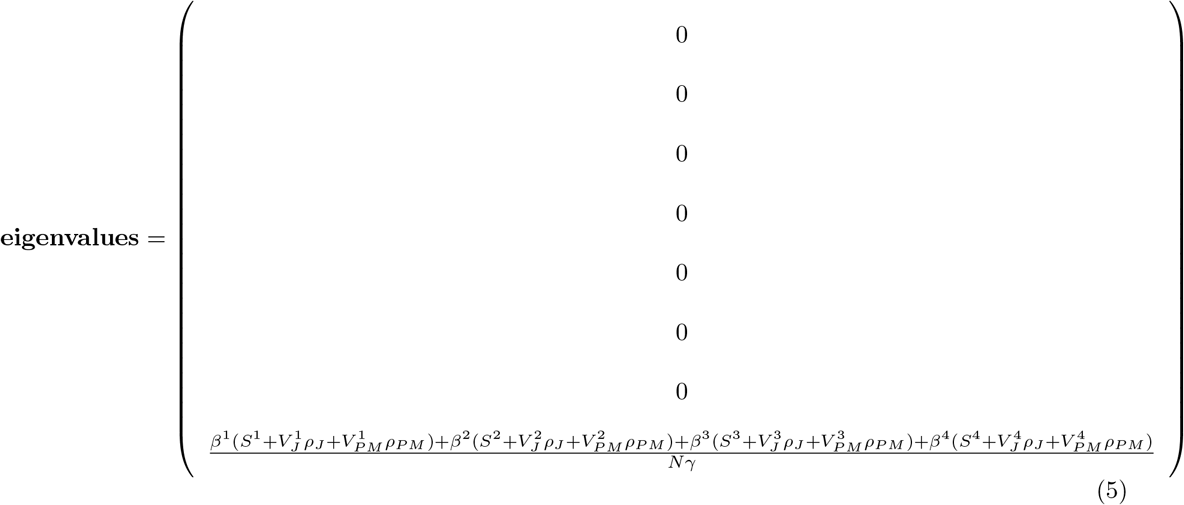

The largest positive eigenvalue of the next generation matrix is the basic reproductive number (R_*effective*_) for the system.

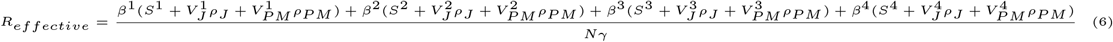

